# Evaluating the Relationship between Neighborhood-Level Social Vulnerability and Patient Adherence to Ophthalmology Appointments

**DOI:** 10.1101/2022.06.22.22276771

**Authors:** Angelica C. Scanzera, Sasha Kravets, Joelle A. Hallak, Hugh Musick, Jerry A. Krishnan, R.V. Paul Chan, Sage J. Kim

## Abstract

**Objective:** Little is known about the association between neighborhood characteristics and non-adherence to attending scheduled ophthalmology appointments. The purpose of this study was to examine the association between neighborhood-level social vulnerability and adherence to scheduled ophthalmology appointments.

**Design:** Retrospective cohort study.

**Participants:** Adults aged 18 years and older with scheduled ophthalmology appointments between September 12, 2020, and February 8, 2021.

**Methods:** A single-center study was conducted at the University of Illinois Chicago Illinois Eye and Ear Infirmary, an urban tertiary care referral center in Chicago, Illinois. Primary exposure is neighborhood-level Social Vulnerability Index (SVI), based on the patient’s address of residence. The SVI ranks (possible range 0 to 1) each census tract on 15 social factors into four related themes (socioeconomic status, household composition & disability, minority status & language, and housing type & transportation). Higher SVI rankings indicate higher levels of social vulnerability. The overall SVI ranking and rankings for each of the four themes were analyzed separately as the primary exposure of interest in multivariable logistic regression models that controlled for age, sex, status (new or established appointment), race, and distance from clinic.

**Main Outcome Measure:** Non-adherence to attending scheduled ophthalmology appointments, defined as missing more than 25% of scheduled appointments.

**Results:** A total of 8,322 unique patients (41% non-Hispanic Black, 24% Hispanic, 22% non-Hispanic White) had scheduled appointments during the five-month study period (range 1 to 23 appointments). Of those, 28% of patients were non-adherent to appointments. In multivariable logistic regression models, non-adherence to appointments was associated with living in higher SVI ranking neighborhoods (socioeconomic status: (adjusted odds ratio [95% confidence interval]) 2.38 [1.94, 2.91]; household composition/disability: 1.51 [1.26, 1.81]; minority status/language: 2.03 [1.55, 2.68]; housing type/transportation: 1.41 [1.16, 1.73]; and overall SVI: 2.46 [1.99, 3.06]).

**Conclusions:** Neighborhood-level measures of social vulnerability are associated with greater risk of non-adherence to scheduled ophthalmology appointments. Studies to better understand these neighborhood-level vulnerabilities are needed to inform the design and evaluation of multi-level (individual and neighborhood) strategies to reduce disparities in access to ophthalmology care.

---

The Healthy People 2030 Initiative has maintained a goal to increase the proportion of adults who have had a comprehensive eye exam in the last 2 years.^1^ Despite strong evidence supporting this initiative, disparities continue to exist. The prevalence of glaucoma, diabetic retinopathy, and overall visual impairment is greater in Blacks and Hispanics compared to non-Hispanic Whites.^2, 3^ In addition to biologic factors, these findings are the result of social determinants of health, ^3-5^ and recognizing the social context contributing to these disparities is critical.

Vision loss is a public health issue disproportionately affecting marginalized communities,^6-11^ It can limit every aspect of an individual’s daily life, including communication, education, independence, mobility, and career goals,^12^ and is one of the most feared disabilities in the United States.^13^ Screening for refractive error and early eye disease could prevent a high proportion of unnecessary vision loss or blindness.^10^ For example, one study found that 50% of subjects receiving an ophthalmologic screening examination had an improvement in vision after refractive correction alone.^14^ However, minorities and people of low socioeconomic status underutilize eye care, are disproportionately affected by barriers to care, and are at the greatest risk of vision loss.^6, 7^

It is widely accepted that an individual’s health is determined primarily by factors outside of the healthcare system, such as poverty, unemployment, or lack of access to care.^15, 16^ In a previous study at our institution early in the COVID-19 pandemic, we found that individuals scheduled for recommended urgent eye appointments who did not adhere more often came from neighborhoods with a greater proportion of Blacks, greater unemployment rates, and a greater number of COVID-19 related deaths.^17^ A higher neighborhood unemployment rate continued to be significantly associated with non-adherence after controlling for race and cumulative deaths from COVID-19, showing the importance of understanding neighborhood context.

Little is understood about the modifiable and non-modifiable neighborhood-level determinants contributing to issues in eye care access. One potential tool to consider in better understanding these neighborhood-level determinants is the Social Vulnerability Index (SVI). Social vulnerability is defined as the potential negative effects on communities caused by external stressors on human health.^18^ Such stressors include natural disasters or health crises such as the ongoing COVID-19 pandemic, which disproportionately affect resource poor communities and contribute to human suffering.^19^ In particular, the Centers for Disease Control and Prevention (CDC) SVI is a composite measure that represents neighborhood relative vulnerability compared to all other communities nationally by census tract. U.S. Census data is used to create a percentile rank for each census tract using 15 social factors organized into 4 themes: socioeconomic status, household composition & disability, minority status & language, and housing type & transportation (**Figure 1**). Though SVI is typically used in response to emergency events or disasters,^20^ it has recently been studied in healthcare to better understand access and outcomes.^21, 22^

**FIGURE 1:**
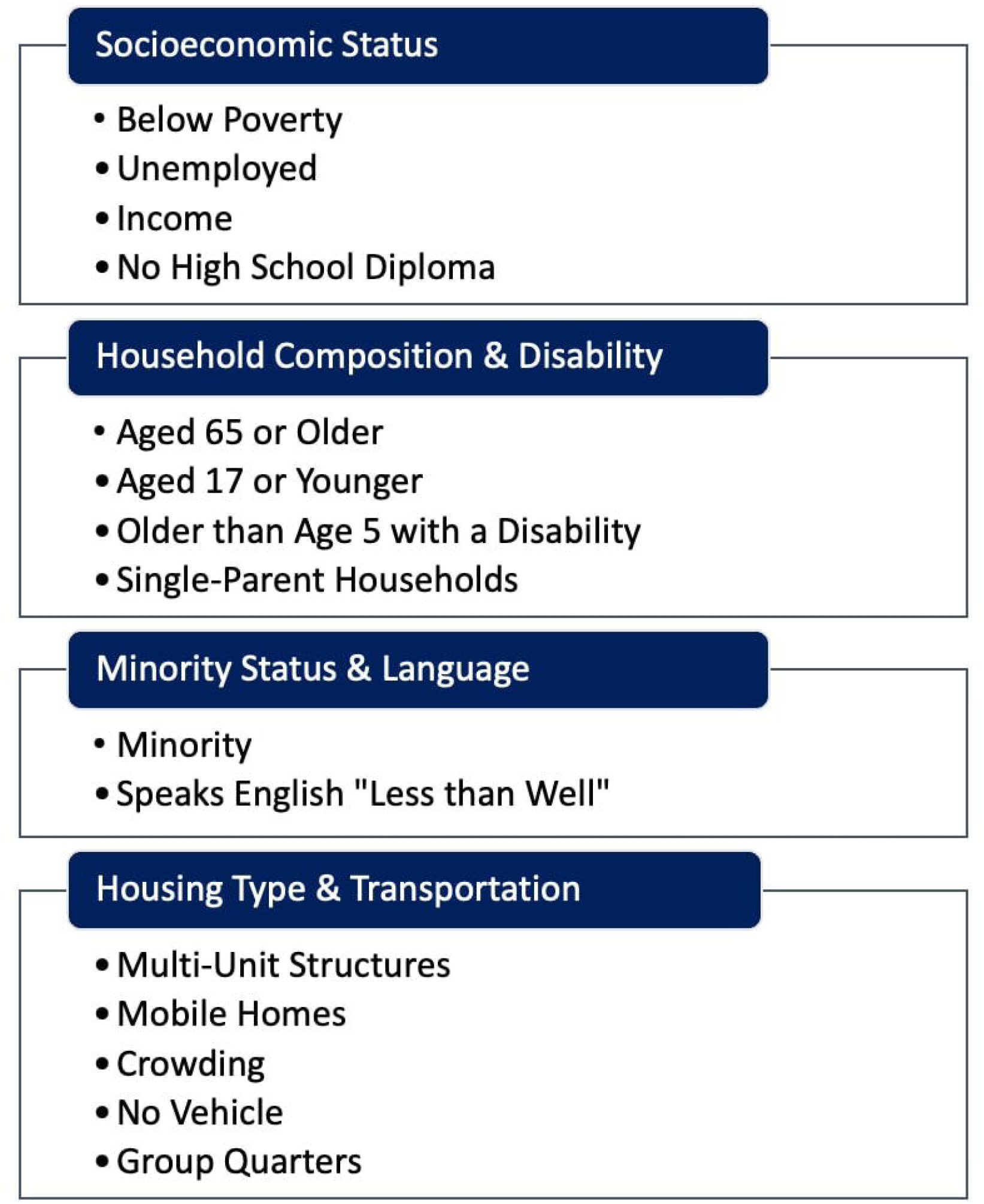
SOCIAL VULNERABILITY INDEX (SVI). Social factors are categorized into 4 themes which make up the overall SVI.

In the interest of improving eye health equity, we utilized SVI as a potentially applicable measure to assess disparities in adherence and target interventions. The objective of this study was to examine the association between neighborhood-level social vulnerability and adherence to scheduled ophthalmology office appointments at an urban tertiary referral center.

## METHODS

### Design, Setting, & Participants

In this retrospective cohort study, we assessed the association between patient individual-level variables and neighborhood-level social vulnerability on adherence to scheduled ophthalmology appointments at an urban tertiary care referral center. All individuals 18 years and older scheduled for an ophthalmology appointment between September 12, 2020, and February 8, 2021, at the Department of Ophthalmology at the University of Illinois Chicago were included in this study. This study was approved by the University of Illinois at Chicago Institutional Review Board (protocol 2021-0177). This research adhered to the tenets of the Declaration of Helsinki.

### Exposure

The primary exposure of interest is neighborhood-level social vulnerability index (SVI) based on the patient’s address of residence. The 2018 Centers for Disease Control and Prevention SVI was used for this study.^18^ Percentile ranking values range from 0 to 1, with higher values indicating greater vulnerability. Each census tract receives a separate percentile ranking for individual themes as well as an overall composite score. Residential addresses were geocoded using ArcGIS, a geographic information systems software, to append U.S. Census community characteristics including overall SVI, SVI of each theme, and each of the 15 social factors by census tract.

Secondary exposures included individual-level variables, which were provided through a medical record review. Variables included age, gender, race/ethnicity, insurance status, distance from the clinic (miles), new patient status, number of appointments scheduled during the study period, and appointment status (reported as completed or no-show in the record). A patient was considered new if they had not previously been evaluated in the service they were scheduled in.

### Outcome

The main outcome of interest was non-adherence to attending scheduled ophthalmology appointments. Patients had varying numbers of scheduled appointments (between 1 and 23). We looked at those who missed 20%, 25%, and 30% of their scheduled appointments, and the proportion of individuals who would be defined as non-adherent was similar (29.6%, 28%, 27.9%). For the purposes of this study, a patient was considered non-adherent if they missed more than 25% of scheduled appointments.

### Statistical Analysis

Mean ± Standard Deviation (SD) or median ± inter-quartile range (IQR) and proportions were reported for continuous and categorical variables, respectively. A t-test or Wilcoxon rank-sum test was conducted to assess a difference in continuous variables by adherence status, and a Chi-square test or Fisher’s exact test was used to assess a difference in categorical variables and adherence status. SVI rankings for each of the four themes, as well as an overall composite ranking, were analyzed separately as the primary exposure of interest in multivariable logistic regression models that controlled for age, sex, status (new or established appointment), race/ethnicity, and distance from clinic. An interaction between race and SVI was initially examined but was found to be insignificant. Using the logistic regression model, the predicted probability of non-adherence was determined for each patient. A p-value of ≤ 0.05 was considered statistically significant. Data were analyzed using R (R Core Team (2019). R: URL https://www.R-project.org/).

## RESULTS

A total of 8,322 unique patients were scheduled for an ophthalmology appointment during the study period. The number of scheduled appointments ranged from 1 to 23 per patient. Just over 1 in 4 (28%) patients were non-adherent. Mean age was 57 ±17 years. Most patients were women (59.2%), and non-Hispanic Black (41.2%), Hispanic (24.1%), or non-Hispanic White (21.9%). Over two-thirds of patients had public insurance (37.2% Medicare, 34.5% Medicaid), and 29.7% were new patients. Median (IQR) distance to the clinic that patients traveled was 7.0 (±10) miles (**Table 1**). Patients scheduled for an ophthalmology appointment come from communities where the mean percentage of persons living below poverty is 20.3%, unemployment rate is at 10.6%, minorities accounted for 68.3%, and per capita income median (IQR) estimate is $24,541 ($18,557) (ranging from $2,530 to $130,543).

**TABLE 1:** INDIVIDUAL CHARACTERISTICS IN PATIENTS SCHEDULED FOR OPHTHALMOLOGY APPOINTMENT BY ADHERENCE

|  | **Overall**<br>n=8,322 | **Adherent**<br>n=5,988 | **Non-adherent**<br>n=2,334 | **P-value** |
| --- | --- | --- | --- | --- |
| **Individual Characteristics** |  |  |  |  |
| Mean Age (years ± SD) | 56.7 ±17.1 | 57.7 ±16.9 | 54.2 ±17.3 | <0.01 |
| Median Distance to Clinic (miles ± IQR) | 7.0±10 | 7.5 ±11.8 | 6.9 ± 9.3 | <0.001 |
|  | n (%) |  |  |  |
| Gender |  |  |  |  |
| Female | 4,924 (59.2) | 3552 (72.1) | 1372 (27.9) | 0.67 |
| Male | 3,397 (40.8) | 2435 (71.7) | 962 (28.3) |  |
| New Patient |  |  |  |  |
| Yes | 2,470 (29.7) | 1675 (67.8) | 795 (32.2) | <0.01 |
| No | 5,852 (70.3) | 4313 (73.7) | 1539 (26.3) |  |
| Race |  |  |  |  |
| Asian | 362 (4.40) | 297 (82.0) | 65 (18.0) | <0.01 |
| Black | 3,429 (41.2) | 2191 (63.9) | 1238 (36.1) |  |
| Hispanic | 2,006 (24.1) | 1398 (69.7) | 608 (30.3) |  |
| White | 1,822 (21.9) | 1586 (87.1) | 236 (13.0) |  |
| Other | 703 (8.5) | 516 (73.4) | 187 (26.6) |  |
| Insurance Type |  |  |  |  |
| Medicaid | 2,871 (34.5) | 1840 (64.1) | 1031 (35.9) | <0.01 |
| Medicare | 3,094 (37.2) | 2334 (75.4) | 760 (24.6) |  |
| Private | 1,957 (23.5) | 1551 (79.3) | 406 (20.8) |  |
| Self-Pay | 264 (3.2) | 144 (54.5) | 120 (45.5) |  |
| Other Public | 114 (1.4) | 98 (86.0) | 16 (14.0) |  |
| Worker's Comp | 22 (0.3) | 21 (95.5) | 1 (4.5) |  |

### Individual-level Determinants

Table 1 summarizes individual characteristics by adherence. There was no significant difference by sex; however, non-adherent patients were slightly younger (54.2 vs. 57.8 years; P<0.01) and living closer (6.92 vs. 7.37 miles; P<0.01). Non-adherence was greatest among Blacks (36.1%), followed by Hispanics (30.3%), other (26.6%), Asian (18.0%), and White (13.0%; P<0.01). New patients were more likely to be non-adherent (32.2%) compared to established patients (26.3%; P<0.01). Non-adherence was also greatest among uninsured patients (45.5%) followed by those with Medicaid (35.9%).

### Neighborhood-level Determinants

Median overall SVI in this population was 0.70. Median SVI was higher in non-adherent (0.78) compared to adherent patients (0.66; P<0.01). In multivariable logistic regression models, non-adherence to appointments was more likely with higher SVI rankings (socioeconomic status: (adjusted odds ratio [95% confidence interval]) 2.38 [1.94, 2.91]; household composition/disability: 1.51 [1.26, 1.81]; minority status/language: 2.03 [1.55, 2.68]; housing type/transportation: 1.41 [1.16, 1.73]; and overall SVI: 2.46 [1.99, 3.06]). **Table 2** provides the multivariable regression model using overall SVI as the primary exposure. Median SVI was greater in non-adherent compared to adherent patients for each of the four individual themes (P<0.01). The greatest difference was noted for socioeconomic status, in which non-adherent patients had a median SVI of 0.82 compared to 0.67 in adherent patients. **Figure 2** further illustrates themed median SVI by adherence.

**TABLE 2:**
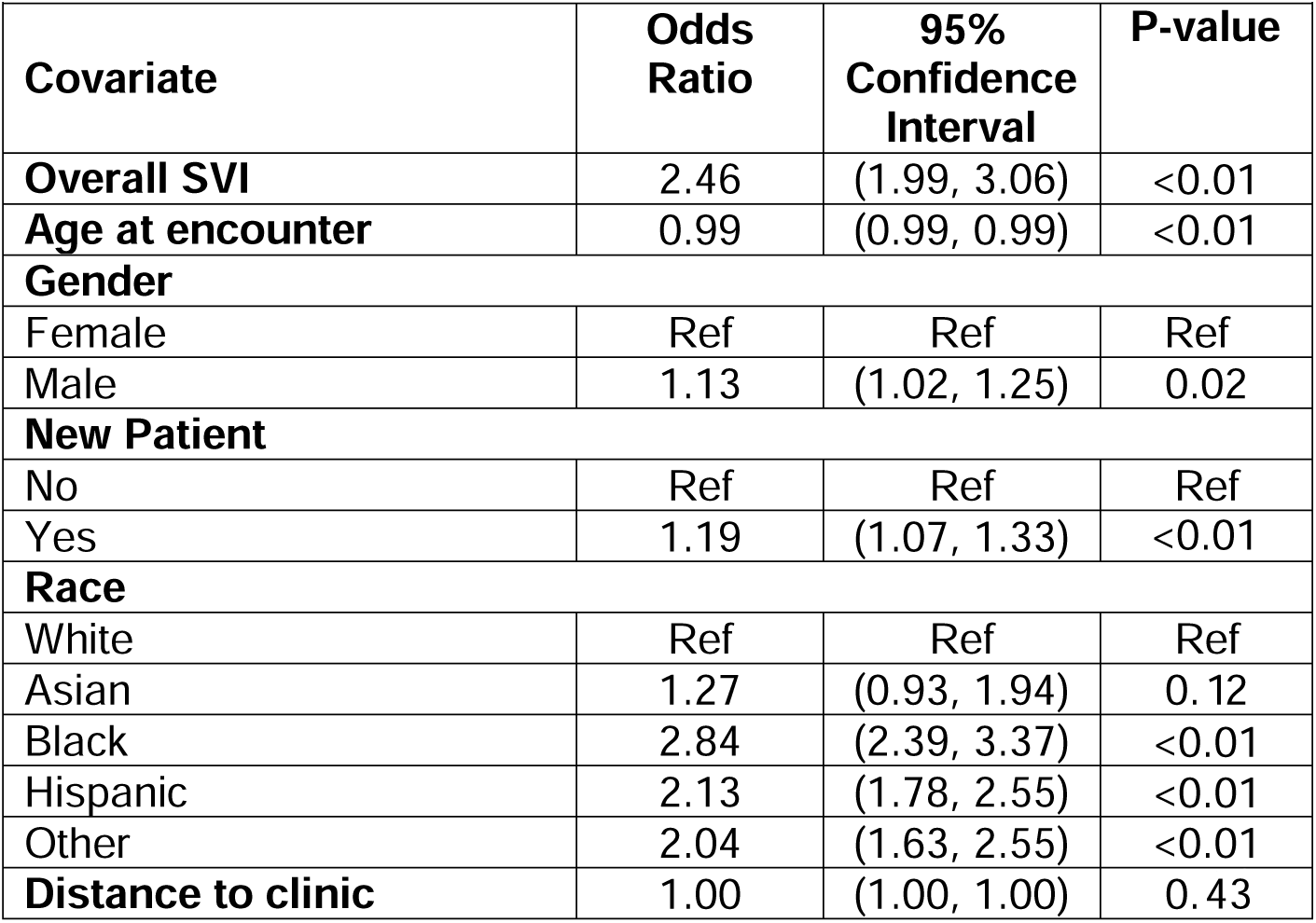
MULTIVARIABLE MODEL OF SOCIAL VULNERABILITY INDEX (SVI) WITH NON-ADHERENCE ADJUSTING FOR INDIVIDUAL LEVEL COVARIATES

**FIGURE 2:**
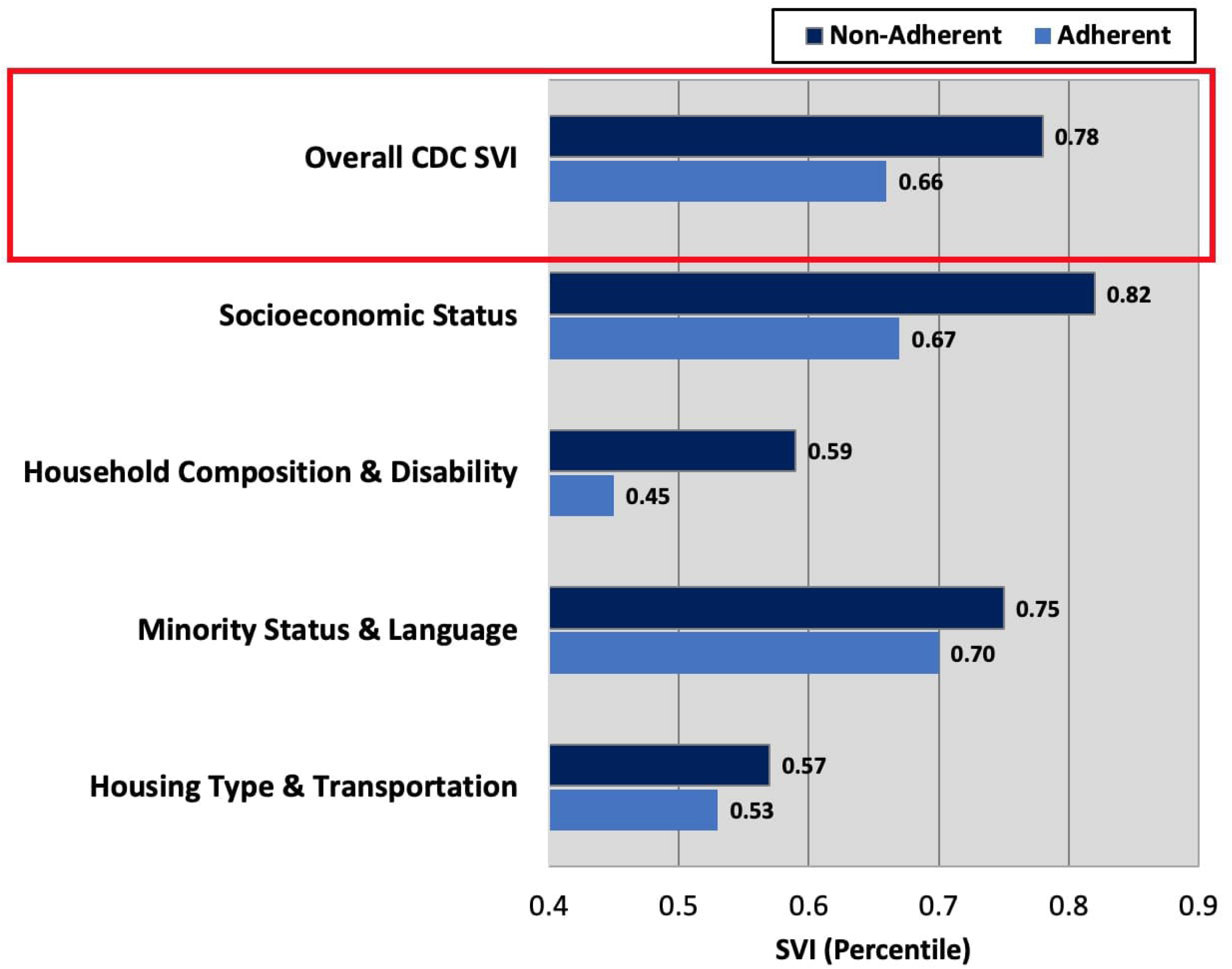
THEMED MEDIAN SOCIAL VULNERABILITY INDEX (SVI) BY APPOINTMENT STATUS.

Median neighborhood SVI was highest among Blacks (0.79) followed by Hispanics (0.77), other (0.57), Asians (0.45), and Whites (0.30). When controlling for age, sex, patient status, distance from clinic and overall SVI, the odds of non-adherence (95% CI) were 2.8 (2.39, 3.37) and 2.1 (1.78, 2.55) for non-Hispanic Blacks and Hispanics, respectively, when compared to non-Hispanic Whites. Using the multivariable model, we computed the predicted probability of non-adherence for a range of SVI from 0-1 for each racial or ethnic group. **Figure 3** illustrates that as neighborhood SVI increases, the probability of non-adherence increases, regardless of a racial category. Additionally, we see that the probability of non-adherence is highest for Blacks, regardless of neighborhood SVI, and increases at a somewhat faster rate, compared to all other races. Specifically, the mean predicted probability of non-adherence for non-Hispanic Blacks is 0.32, compared to 0.14 for non-Hispanic Whites.

**FIGURE 3:**
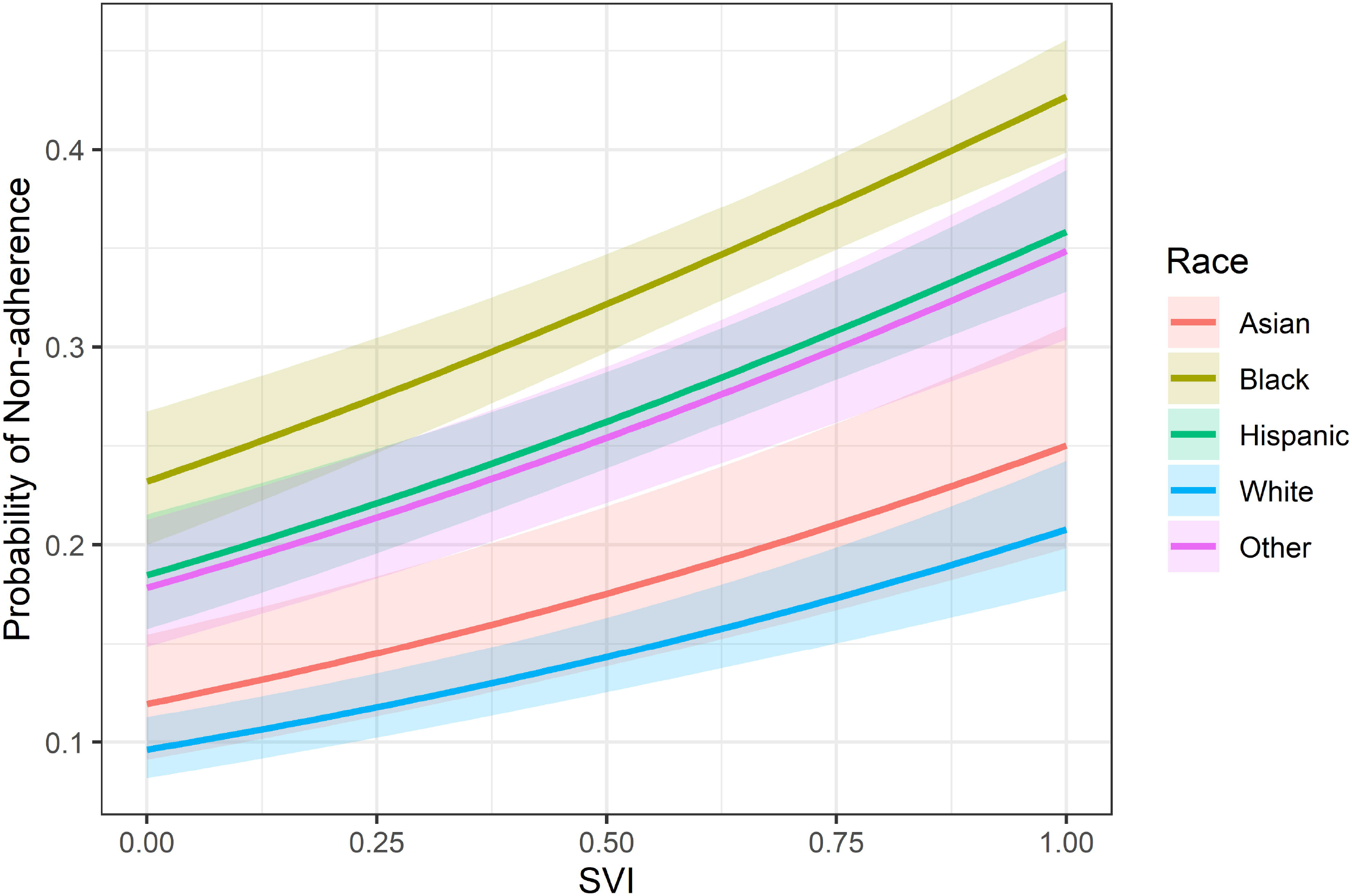
PROBABILITY OF NON-ADHERENCE USING SOCIAL VULNERABILITY INDEX (SVI). Looking at each race individually, as SVI increases, the probability of non-adherence increases, adjusting for all other variables.

## DISCUSSION

We evaluated the effects of individual and neighborhood-level factors on adherence to scheduled ophthalmology appointments at an urban tertiary referral center. The key findings of this study are: 1) greater neighborhood-level social vulnerability is associated with a greater likelihood of non-adherence to ophthalmology appointments, and 2) after adjusting for neighborhood-level vulnerability and other relevant factors, race/ethnicity continued to be significantly associated with non-adherence to appointments.

We demonstrated that CDC SVI is a useful tool to examine disparities in adherence and outcomes within ophthalmology. Though CDC SVI was established to assist with understanding vulnerability to disasters, this study demonstrates the use of SVI to examine health disparities, particularly in ophthalmology appointment adherence. To the authors’ knowledge, this is the first study that assesses the use of the CDC’s SVI in ophthalmology. However, SVI has been widely used in health science research. For example, higher neighborhood SVI has been associated with an increased number of chronic conditions,^23^ worse surgical outcomes,^21^ and increased risk of mortality.^24^ The use of SVI to understand the patient’s neighborhood context expands social determinants of health outcomes in ophthalmic research.

Overall, greater neighborhood-level SVI is associated with a greater likelihood of non-adherence to ophthalmology appointments, and the largest difference in adherence was observed with the socioeconomic status SVI theme. Low socioeconomic status is associated with a higher incidence of open globe injury,^25^ presenting at a more advanced stage of age-related macular degeneration,^26^ a greater risk of developing glaucoma,^27^ and higher rates of blindness.^28^ Residency in more disadvantaged neighborhoods is associated with poor adherence to both diabetic retinopathy screening^29, 30^ and physician recommended urgent ophthalmology appointments.^17^ These findings indicate the importance of understanding the communities our patients reside in. Moving forward, SVI is being integrated into many electronic health records, which opens the opportunity for incorporating social context into the standard of care and clinical interventions.

While non-adherence was higher in racial/ethnic minorities, there was also a stark difference in neighborhood-level vulnerability by race/ethnicity in our patient population. Median neighborhood-level SVI in Blacks and Hispanics were significantly higher than in Whites (0.79, 0.77, and 0.30, respectively). Neighborhood level inequities are well recognized and exacerbated by residential segregation. Chicago is a highly segregated city, with a dissimilarity index over 56, meaning that 56% of Chicago’s population would have to move to another neighborhood to balance the composition of individual neighborhoods to the region’s general demographic composition.^31^ Racial residential segregation affects how neighborhood investment decisions are made which has resulted in uneven access to health care ^32, 33^ and consequently, shorter life expectancies in highly segregated Black communities.^34, 35^ Racial minorities are more likely to live in underserved areas with lower levels of health and social service opportunities, further perpetuating poor health outcomes.^36-39^ The National Institute on Minority Health and Health Disparities (NIMHD) Research Framework describes a multidimensional model that depicts a comprehensive set of health determinants which includes domains of influence over the life course (biological, behavioral, physical/built environment, sociocultural environment, and the health care system) and levels of influence (individual, interpersonal, community, and societal).^40^ The intersections of the domains with each level of influence have distinct effects on health outcomes. Community and societal investment could improve individual behavior, in our case, adherence to appointments, and health outcomes.

Interestingly, after adjusting for neighborhood-level SVI and other relevant factors, Blacks and Hispanics continued to have greater odds of non-adherence to scheduled ophthalmology appointments compared to Whites, indicating that other neighborhood and individual-level factors are at work, beyond SVI. This is consistent with a study of almost 80,000 Medicare beneficiaries with glaucoma which found that eye care utilization disparities were greatest among Blacks and Hispanics, even after stratifying by socioeconomic status.^41^ One hypothesis for this difference focuses on concentrated affluence rather than disadvantage. Brooks-Gunn et al argued that concentrated socioeconomic resources or affluence, not captured by SVI, can have a positive influence even after controlling for individual family resources.^42^ While individual-level social capital has the greatest influence on health, ^43, 44^ increased neighborhood-level social capital is also positively related to better health.^43-46^ in our patient population, minorities live in more disadvantaged areas while Whites live in less disadvantaged areas. This spatial inequality results in social, economic, and political isolation,^47^ which could explain the differences in eye care utilization in racial and ethnic minorities found in our study not captured with SVI.

Another possibility is that racial discrimination could be a deterrent to medical care. Racial discrimination is a risk factor for disease and a contributor to structural racism in health care.^48^ Findings from a large meta-analysis found that individuals who reported experiences of racial discrimination were two to three times as likely to be less trusting of health care workers and systems, perceive lower quality of and satisfaction with care, and express less satisfaction with patient-provider communication and relationships. Experiencing racism was also associated with delays in seeking health care and reduced adherence to medical recommendations.^49^ Within eye care, lack of trust, empathy, or patient-doctor communication have each been emphasized as barriers to utilization in populations at high risk of vision loss,^6^ and minorities have reported feeling less respected by health care professionals compared to non-Hispanic Whites.^50^ There have been several calls to diversify the workforce to improve patient care and focus on improving health equity through research^51, 52^ as both ophthalmology and the field of medicine currently lack diversity.^53^ Given these findings, it is imperative for providers to acknowledge that structural racism exists in healthcare, recognize and work toward ways to address implicit bias, and diversify the workforce.

Our study has several limitations that should be considered. 1) Most notably, this study investigated data at a single center. Given that our patient population is more diverse than the make-up of the city of Chicago, this may limit the generalizability of our study; however, these findings are most relevant to more vulnerable populations. Future multi center studies are needed to confirm these findings. 2) We also focused on demographic factors and did not leverage all variables available within the medical record. The patient’s ocular diagnosis and comorbidities should be considered in future studies to consider interaction or confounding variables. 3) This study used 2018 CDC SVI, the latest available SVI, which was data reported prior to the pandemic. Neighborhood factors have likely changed since the onset of the pandemic. We imagine disparities may be even greater than what is reported. 4) Lastly, an important consideration is that neighborhood-level SVI helps us to understand the community our patients are coming from but cannot be easily applied to the individual. This data suggests the use of SVI to identify patients at higher risk of non-adherence; however, interventions should be considered at the patient, health-system, community, and policy levels.

Previous reports of ambulatory care no-show risk prediction models have focused primarily on hospital components and individual behavior, such as appointment lead time, appointment rescheduled by the provider, or history of previous no-show;^54, 55^ These fail to take social context into account. Further, interventions focus on clinical productivity, suggesting options such as added reminder calls or text messages to improve adherence,^56^ or creating overbook slots, which fail to address factors driving no-shows and likely intensify health inequities. Findings from this study help us understand who is less likely to show for an appointment. Utilizing variables that account for social context, there is potential to identify patients at higher risk of no-show and provide them with targeted interventions.

This study serves as a step in preintervention planning.^57^ Participatory science elevates patient and community voices and is essential to gain insight into how our patients interact with the healthcare system.^58^ In order to generate changes in healthcare delivery to promote health equity, a critical next step is to engage stakeholders, including patients and providers, to identify the barriers and facilitators that exist after scheduling an eye appointment that prevent a patient from adhering to this appointment.

In this study, we examined the association between neighborhood-level social vulnerability and adherence to scheduled ophthalmology appointments at an urban tertiary care referral center. We found that higher neighborhood-level social vulnerability is associated with an increased risk of non-adherence to ophthalmology appointments. Poor adherence to appointments is not entirely attributed to SVI in minorities, as race/ethnicity continued to be significantly associated with non-adherence to scheduled appointments after controlling for other relevant factors. These findings suggest the potential effects of individual and neighborhood-level determinants on non-adherence to scheduled ophthalmology appointments. Studies to inform the design and evaluation of multi-level (individual and neighborhood) strategies to reduce disparities in access to ophthalmology care are needed.

## Data Availability

All data produced in the present study are available upon reasonable request to the authors

## Acknowledgments

The authors thank the Center for Health Equity Research (CHER) Chicago for their invaluable collaboration.

## Acronyms

SVI: Social Vulnerability Index
SD: Standard Deviation
IQR: Inter-Quartile Range

